# Precision Multiparameter Tracking of Inflammation on Timescales of Hours to Years Using Serial Dried Blood Spots

**DOI:** 10.1101/2019.12.09.19014233

**Authors:** N. Leigh Anderson, Morteza Razavi, Matthew E. Pope, Richard Yip, L. C. Cameron, Adriana Bassini-Cameron, Terry W. Pearson

## Abstract

High-frequency longitudinal tracking of inflammation using dried blood microsamples provides a new window for personalized monitoring of infections, chronic inflammatory disease, and clinical trials of anti-inflammatory drugs. Using 1,662 dried blood spot samples collected by 16 subjects over periods of weeks to years, we studied the behavior of 12 acute phase response (APR) and related proteins in inflammation events correlated with infection, vaccination, surgery, intense exercise and Crohn’s disease. Proteins were measured using SISCAPA mass spectrometry and normalized to constant plasma volume using low-variance proteins, generating high precision within-person biomarker trajectories with well-characterized personal baselines. The results shed new light on the dynamic regulation of APR proteins and offer a new approach to visualization of multidimensional inflammation trajectories.

## Introduction

In this paper we explore the longitudinal behavior of inflammation-related acute phase response (APR^1^) proteins in a unique collection of dried blood spot (DBS) samples. These included 1,522 samples collected longitudinally by eight individuals over periods of up to nine years (with extended periods of daily sampling) and 140 samples collected by eight elite athletes at multiple time points per day during one week of Olympic training using a Sportomics approach [1]. Using peptide-based SISCAPA immunoaffinity-mass spectrometry (SISCAPA-MS) and a novel optimized plasma volume normalization method, we measured the scale and temporal relationships between inflammatory proteins and the frequencies of large and small changes from personal baselines. We also developed novel means for visualizing this rich data.

Inflammation is central to the understanding of human health and disease. Underlying the classical definition (heat, pain, redness, and swelling), inflammation involves a complex balance between protective and destructive processes implemented by a varied cast of proteins under active regulatory control. In clinical contexts, these complex processes are usually summarized by measuring only a single blood biomarker: C-reactive protein (CRP [2]), a key component of the innate immune system involved in recognizing and destroying bacterial pathogens. Enormous clinical value has been obtained through quantitative measurement of CRP, as reflected by the fact that there are more FDA-cleared commercial tests for CRP than for any of the >100 other clinically measured blood proteins. CRP levels have well-established clinical significance over a very wide dynamic range, from small (less than 2-fold) increases associated with increased cardiovascular disease risk [3] to increases of more than 100-fold in major infections [4]. Considered more broadly, increases in CRP are associated with negative developments in a wide range of health situations, including infection [5], arthritis [6], surgery [7], intense exercise [8], sleep apnea [9], depression [10], air pollution [11], welding fume exposure [12], Parkinson’s disease [13], pregnancy [14], inflammatory bowel disease [15] and various cancers [16], to name a few examples.

Many therapeutics that reduce inflammation are in use or under development. These usually act either directly (by targeting inflammation pathway signaling) or indirectly (by treating causes of inflammation). Inflammation signaling is generally targeted by biologics, including a wide and rapidly growing range of molecules such as the anti-TNF alpha antibodies infliximab [17] and adalimumab [18]; TNF inhibitor etanercept [19]; anti-IL-6 mAbs siltuximab [20] and olokizumab [21]); anti-IL-6 receptor tocilizumab [22]; anti-IL-1β mAb canakinumab [23]; and IL-1 receptor antagonist anakinra [24]. ISIS 329993, an antisense oligonucleotide complementary to the coding region of the human CRP messenger RNA, directly targets CRP translation [25]. A similarly wide spectrum of small molecules target causes of inflammation, including antibiotics (such as teicoplanin [26], vancomycin [27] and cefuroxime [28]), various statins [29], the COX/5-LOX inhibitor tenidap [30], methotrexate [31], the Lp-PLA2 inhibitor darapladib [32], and the MAPK inhibitor dilmapimod [33]. All of these drugs reduce CRP levels, and in fact CRP has been used as a pharmacodynamic biomarker in trials of many of them as a means of selecting dose and dosing schedule [18,19,21–23,31]. CRP thus carries a heavy load in measuring inflammation, both in terms of diagnostic evaluation and in the assessment of treatment efficacy.

Can a single biomarker adequately summarize such complex mechanisms and outcomes? The current use of CRP as a biomarker implicitly assumes that inflammation can be considered a fixed response [34] to many disparate triggers, following the same basic script in all individuals.

In reality, the inflammatory response exhibits wide variations in scope and timing, as well as major differences between individuals. At the molecular level, it involves significant changes in the concentrations of many proteins besides CRP, including cytokines (short-lived, low-abundance signaling proteins that regulate inflammatory responses [35]) and a broad set of acute phase response (APR) proteins [36] that implement many of the functions required to recognize and deal with inflammatory stimuli. The cytokines, including primary inflammation regulators such as interleukin-6 (IL-6), are of central importance in research on regulatory mechanisms, but are only occasionally used as clinical diagnostic tests because of their short half-lives, localized pleotropic effects and very low concentrations (low pg/mL). APR proteins like CRP, on the other hand, represent the systemic effects of cytokine regulation and have longer half-lives, a wide range of specific effector functions and higher concentrations. Differences among APR proteins in terms of their regulation, half-lives and functions imply that each has the potential to contribute additional non-redundant diagnostic information, which in turn suggests that a panel of APR proteins would have greater sensitivity and power than CRP alone in analyzing inflammatory responses. In this paper we explore that concept using a set of twelve APR and APR-related proteins in blood, each with distinct roles and behaviors. These include clinically-established molecules involved in both innate (SAA, CRP, LPSBP, MBL; see Table 2 for complete list of protein abbreviations) and adaptive (IgM) immune responses, as well as proteins involved in drug transport (Alb, A1AG), iron scavenging and transport (Hp, Hx), the complement cascade (C3), coagulation (FibG) and neutrophil activity (MPO). High precision measurement of protein panels in DBS is achieved by mass spectrometry combined with a sample preparation workflow involving stable isotope standards and capture by anti-peptide antibodies (SISCAPA [37–39]). From an informatics viewpoint, this breadth of analysis increases the functional dimensionality of inflammation measurement, giving a clearer picture of what is happening at any moment in time.

**Table 1.**
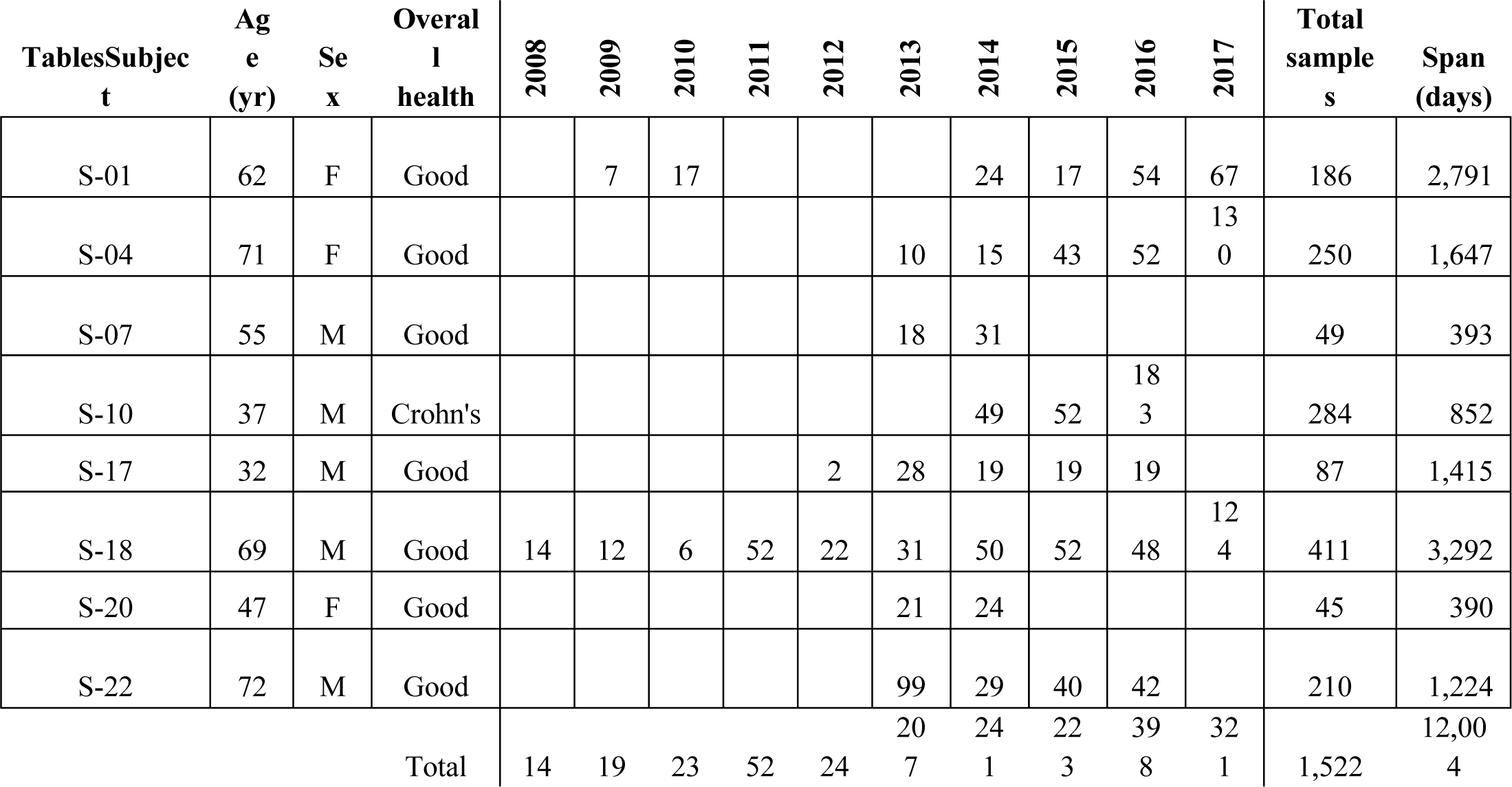
Set I capillary blood DBS samples

**Table 2.** Proteotypic peptides and their proteins measured by SISCAPA-MRM

| Protein | Abbreviation | Role | Peptide | I |  |  |  | II |
| --- | --- | --- | --- | --- | --- | --- | --- | --- |
|  |  |  |  | A | B | C 1 | C 2 | D |
| α-1-acid glycoprotein | A1AG | APR+ | NWGLSVYADKPETTK | 1 | | 1 | | 1 |
| Albumin | Alb | Normalization, APR- | LVNEVTEFAK | 1 | 1 | 1 | 1 | 1 |
| Complement C3 | C3 | Complement, APR+ | IHWESASLLR | 1 |  | 1 |  | 1 |
| C-reactive protein | CRP | Innate immunity, APR+ | ESDTSYVSLK | 1 | 1 | 1 |  | 1 |
| Fibrinogen $\gamma$ chain | FibG | Coagulation, APR+ | YEASILTHDSSIR | 1 | | | 1 | 1 |
| Haptoglobin | Hp | Fe metabolism, APR+ | VTSIQDWVQK | 1 | 1 | 1 |  | 1 |
| Hemopexin | Hx | Fe metabolism, APR+ | NFPSPVDAAFR | 1 | 1 | 1 | 1 | 1 |
| Immunoglobulin M | IgM | Immune response, APR+/- | YAATSQVLLPSK | 1 | 1 | 1 | 1 | 1 |
| LPS-binding protein | LPSBP | Innate immunity, APR+ | LAEGFPLPLLK | 1 | 1 | 1 |  | 1 |
| Mannose binding lectin | MBL | Innate immunity, APR+ | EEAFLGITDEK | 1 | 1 | 1 |  | 1 |
| Myeloperoxidase | MPO | Neutrophil count | DYLPLVLGPTAMR | 1 | 1 | 1 |  | 1 |
| Serum amyloid A | SAA | Innate immunity, APR+ | GPGGVWAAEAISDAR | 1 | 1 | 1 |  | 1 |

Tracking biomarker changes through time completes the biological picture. Concentrations of various inflammation markers change over timescales ranging from minutes to years, exposing associations with a variety of underlying time-dependent disease mechanisms and contextual variables. For this reason, longitudinal studies of serial blood (plasma, sera) samples provide the clearest path to detailed understanding of the dynamics of inflammatory processes needed to distinguish specific causes, guide therapy, predict outcomes and account for differences between individuals. In medicine, longitudinal sampling allows interpretation of results against personal baselines for each protein, thereby personalizing diagnostic interpretation. Such an approach exposes relationships between biomarkers over time, potentially uncovering novel diagnostic multi-parameter indices. In pharmaceutical trials, longitudinal sampling allows construction of mathematical models relating the changing levels of a drug *in vivo* (pharmacokinetics; PK) to changes in one or more biomarkers of effect (pharmacodynamics (PD); together referred to as PK/PD models). However, since frequent longitudinal blood sample collection by venipuncture is generally impractical outside a medical environment (and generally discouraged for ethical reasons, including the associated reduction of subject hematocrit [40]), longitudinal sample collections rarely include more than a 5-10 samples per individual. Thus high-frequency (e.g., daily) sampling is rarely attempted, even in clinical trials.

Longitudinal sample collection can be radically improved by using a less intrusive means of blood collection: fingerprick capillary blood dried on filter paper (“ dried blood spots”: DBS). DBS have been used as clinical samples since the pioneering studies of Guthrie [41] on inborn errors of metabolism in neonates. As practiced routinely by diabetic patients, individuals can collect capillary blood by lancet fingerprick several times each day for glucose measurement without injury or undue discomfort, blood that can be collected and stored in the form of DBS by subjects themselves. Numerous efforts are now underway to further improve the user-friendliness and reproducibility of capillary blood collection that will further enable the approach described here.

Here we report results that illustrate the complex and informative behavior of APR proteins (and related proteins) in tracking inflammation due to multiple causes. The data demonstrate the potential for major improvements in diagnostic test interpretation using personalized baseline values and the feasibility of personalized multiparameter biomarker response models useful in clinical trials.

## Results

Amounts of selected APR and related proteins were measured using an automated, multiplexed SISCAPA-MS protocol [37,39] and normalized to a consistent plasma volume using a personalized approach based on a combined index of three proteins (Alb, Hx, IgM) whose levels are generally very stable in an individual over time (i.e., they exhibit low within-person variance [39]). The calculated scale factors applied to individual longitudinal DBS spots in this study fell in the range of 0.7-1.3 as expected. This volume normalization substantially reduces longitudinal baseline “ noise” in measurements from an individual: Supplementary Figure 1A shows DBS protein levels before and after normalization in 250 serial samples collected by subject S-04 over almost 5 years.

In addition to maintaining stable levels over time, two of the normalizing proteins (Hx and IgM) exhibit large quantitative baseline differences between individuals. As a result, each subject’s sample set, in many cases including hundreds of samples collected over many years, forms a tight cluster on a plot of Hx vs IgM, almost completely separated from the other subjects (Figure 1A). In general, points diverging from the cluster centroids are associated with major inflammation events. Subjects S-04 and S-18 (of opposite sex, contributing 411 and 250 samples respectively) share a living environment, while their sample sets appear as completely separated clusters, indicating that most of this between-subject variation is not determined by environment at the time of collection. Most of the other APR proteins measured showed significant inter-individual differences as well (Supplementary Figure 2).

**Figure 1:**
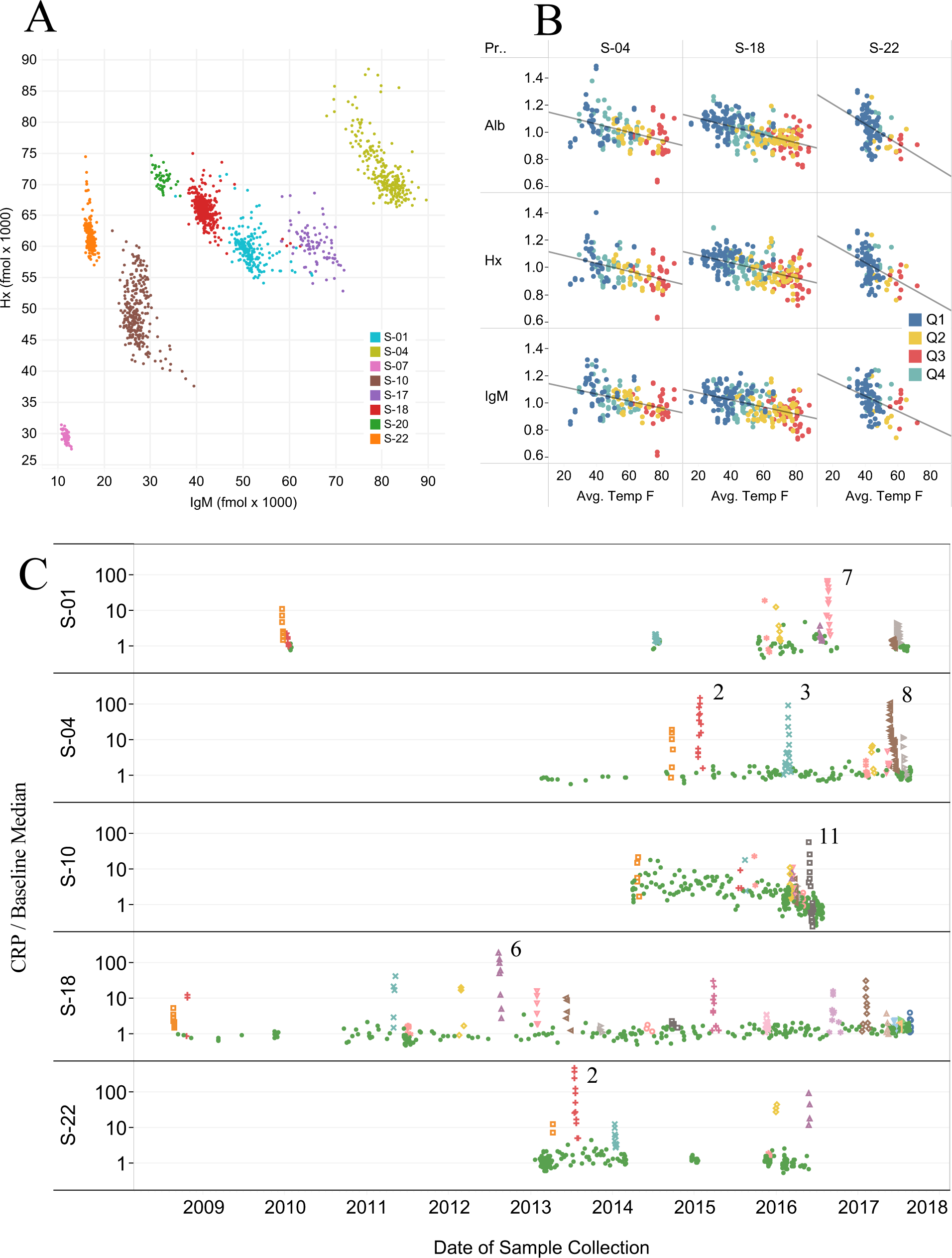
A) Plot of fmol IgM vs Hx for all 1,522 samples in Set I, color coded by subject. B) Abundances of Alb, Hx and IgM (un-normalized fmol divided by personal median value, excluding inflammation event samples) in 3 subjects vs local daily average ambient temperature on the date of collection, colored by calendar quarters. C) Abundance of CRP (normalized fmol divided by personal baseline average) in 5 subjects over time. Inflammation events are color coded (remaining samples are shown as green dots) and most intense events numbered.

While the selected normalizing proteins exhibit striking longitudinal stability, we nevertheless observed two significant sources of variation in these proteins within subjects. Figure 1B shows small (+/- 10%) systematic changes in three proteins in three subjects that correlate with average ambient temperature on the date and location where the samples were collected, an effect previously noted in the form of seasonal variation in some DBS analytes [39]. In addition, as shown in Supplementary Figure 1B, extreme inflammatory events such as major infections affect the concentrations of most plasma proteins to some degree, including those used here for normalization. With the present approach, normalization bias due to inflammation is mitigated due to the fact that while IgM varies little, Alb shows a weak negative APR and Hx shows a weak positive APR, so that the combined index is to a great extent balanced with respect to inflammation.

### Inflammation events

The largest perturbations observed in levels of most proteins occurred during inflammatory events that were noted by the study subjects and were coincident with increases in the acute phase reactants CRP (Fig. 1C) and SAA. However, many events were observed that appear to be “ sub-clinical”: in the five subjects for which extended (>150) longitudinal sample series were available, we identified 57 inflammation events in which CRP was increased significantly above baseline levels in multiple samples collected over a brief interval, allowing identification of a temporal maximum (colored spikes in the CRP time series presented in Fig. 1C). Of these events, approximately half (29 events) included at least one sample in which CRP was increased by more than 10-fold above baseline and all events were confirmed by coordinated increases in CRP, SAA and LPSBP. In contrast to the large increases seen in SAA and CRP in these events, the remaining APR proteins (LPSBP, A1AG, Hp, MBL, FibG, C3 and Hx) showed much smaller effects (Supplementary Figure 1B), with fold-increases relative to baseline ranging from 0.036 (LPSBP) as great as the fold-increase in CRP to 0.002 (Hx).

Given the significant fraction of samples that showed evidence of an inflammatory response, we defined personal “ normal” baseline levels for each biomarker protein based on the subset of samples with CRP below a cutoff equal to the personal median CRP value across all of a subject’s samples; i.e., selecting the half of each subject’s samples with lowest CRP values. For each protein, the median value in these low-inflammation baseline samples was taken as the subject’s personal baseline. The standard deviation in these baseline samples allowed calculation of a personal baseline CV. The average baseline CV over all subjects, samples and proteins was 12.3% (12.4%, 17.0% and 10.2% in Datasets A, B, and C respectively) compared to 38.0% (39.2%, 68.7% and 31.3% in Datasets A, B, and C) for combined baseline and non-baseline (i.e., all) samples (Supplementary Table 3). On average, the ratio of assay CV (from replicate standards) to average subject baseline CV (from subjects’ longitudinal samples) was 0.52, satisfying the criterion (<0.6) allowing statistically meaningful analysis of within-subject biological variation [42] at very low levels of inflammation. In subject S-22, CRP and SAA showed maximum increases of up to 497- and 1,062-fold (equivalent to 2,673 and 3,688 personal standard deviations) respectively, above personal baselines in a major kidney infection, illustrating the >1,000-fold dynamic range of statistically significant (>2 SD) within-person changes. The proportion of samples in which CRP was more than 2-fold higher than the personal baseline ranged from 12-45% among these 5 subjects (Supplementary Table 2).

### Infection

Infections of various kinds constituted the major driver of inflammation events. Figure 2A shows time course plots for 7 significant infections occurring in 5 subjects, each involving SAA increases >100-fold from personal baseline median values. These events were reported by subjects as: respiratory infection (S-01 E7; i.e., event 7 in subject S-01); respiratory infections (S-04 E2, E3 and E8); food poisoning (S-10 E11); pneumonia confirmed by X-ray (S-18 E6); and kidney infection (S-22 E2). Subject S-18, with the largest sample series, exhibited a number of discrete events (Fig. 1C) in addition to S-18 E-6 that were subject-annotated as “ nose colds” and have a remarkably consistent magnitude and structure, likely indicating a reproducible response to similar infectious agents (e.g., rhinoviruses). None of the infections required hospitalization, though in some cases antibiotics were administered.

**Figure 2:**
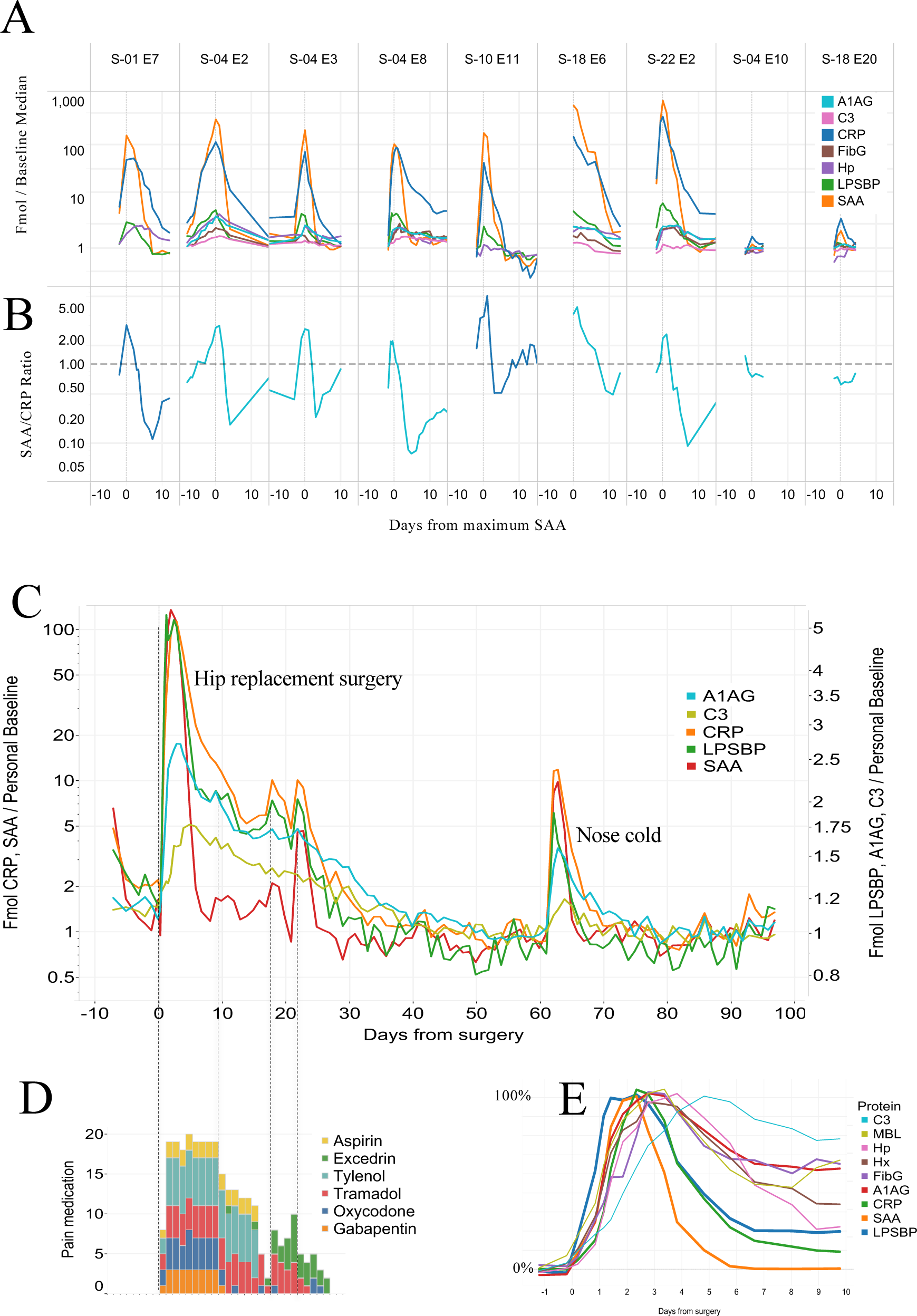
A) Amounts of 6 proteins (normalized fmol divided by personal baseline median, log scale) in 7 largest inflammation events and 2 influenza vaccinations. B) SAA/CRP ratio for the same events shown in A. C) Amounts of 5 inflammation proteins (normalized fmol divided by personal baseline median, two separate log scales) over a 100 day period including a total hip artheroplasty (day 0) and an upper respiratory infection (day 63). D) Requirement for pain medication associated with the surgery. E) Amounts of 9 inflammation proteins over 10 days including a total hip artheroplasty, with each protein normalized to approximately the same scale of change in order to illustrate differences in timecourse.

The ratio of SAA over CRP (each in terms of fold-change from personal baseline median; Figure 2B) shows large changes through the course of each infection. SAA induction exceeds CRP induction at the peak of each event, while it appears that CRP exceeds SAA before and after the peak.

### Influenza vaccination

Two subjects collected daily samples in periods that included a vaccination against influenza (Fluzone™ high dose 2017). Inflammatory responses (Figure 2A; S-04 E10 and S-18 E20) in SAA and CRP were measurable (1.3 – and 2.2-fold increases in SAA from respective personal baselines), but were ∼100 x smaller than those associated with major infections. Of the 57 identified inflammation events, 41 involved SAA increases larger than 2.2-fold (i.e., a greater response than seen with either of these 2 vaccination events).

### Surgery

Subject S-04 underwent elective anterior total hip arthroplasty involving a short (∼2 hr) surgical procedure and subsequent recovery period of 97 days during which daily (or more frequent) samples were collected (Figure 2C). Following an initial rapid rise after surgery, inflammation markers (here plotted on two log scales, left and right, differing by 20-fold in magnitude), declined at different rates until day 55, at which point the levels approached a new baseline approximately 50% below the pre-surgery level. CRP and SAA were induced much more strongly than other inflammation indicators (with maximum inductions of 113- and 136-fold respectively). The magnitudes of the observed responses (in fold-change from personal baselines) were: SAA > CRP >> LPSBP >FibG, Hp, A1AG > MBL, C3 > Hx (136, 113, 5.4, 3.0, 2.9, 2.7, 2.0, 1.8, and 1.2-fold respectively), overall a ∼500-fold range of response magnitudes relative to personal baselines. In the period between days 8 and 30, a series of post-surgical jumps in SAA and CRP (e.g., days 9, 18 and 22) indicated renewed inflammatory activity that generally coincided with increased requirement for pain medication (Figure 2D) and subject reports of strains in the surgical area, followed by smooth declines to baseline. Figure 2C also includes an infection event (an upper respiratory tract infection perceived by the subject as a “ nose cold”) on day 62 following surgery. While the overall magnitude of the inflammation response in this infection was roughly 10% as great as the response to surgery, the kinetics of the early response of the 5 proteins shown were similar.

Figure 2E shows the time course after surgery of nine acute phase proteins normalized to the same peak response (0-100%) and smoothed in order to reveal relative peak times. LPSBP achieved its peak level first (∼1.2 days post-surgery), followed by SAA, CRP, A1AG and MBL, Hp, FibG, Hx and C3, with the last peaking approximately 5 days post-surgery.

### Crohn’s disease

A combination of rapid APR protein changes superimposed on a slow declining trajectory were observed in subject S-10, who has Crohn’s disease and has diligently explored dietary and other measures to control symptoms. Over a period of ∼2.5 yr, CRP and SAA levels were substantially elevated on numerous occasions (Figure 3A shows the entire time period consisting of almost 2 yrs of weekly samples followed by 180 daily samples shown expanded in Fig 3B). Overall, CRP was above 2 times the median personal baseline value in 48% of the individual’s samples, a significantly higher proportion than exhibited by the other subjects. The subject made careful notes of perceived Crohn’s attacks (reported on a scale of 0.5-2 in intensity at times indicated in Fig 3); “ colds” (labeled “ C”) and one episode attributed to food poisoning (“ FP”). It is noteworthy that three of eight recorded Crohn’s attacks occurred at times of lowest CRP (150, 460 and 800 days from first sample), while the rest coincide with CRP peaks. Conversely, at least eight CRP peaks of magnitude equal to those linked to Crohn’s attacks are not noted by the subject as either Crohn’s attacks or infections. Over the period of sample collection, S-10 reported success in significantly reducing the frequency and intensity of gut inflammatory events and this was reflected in the significant downward trend in CRP and SAA, reduced intensity of CRP spikes and a progressive decline of almost 3-fold in Hp, a slowly-responding acute phase protein.

**Figure 3:**
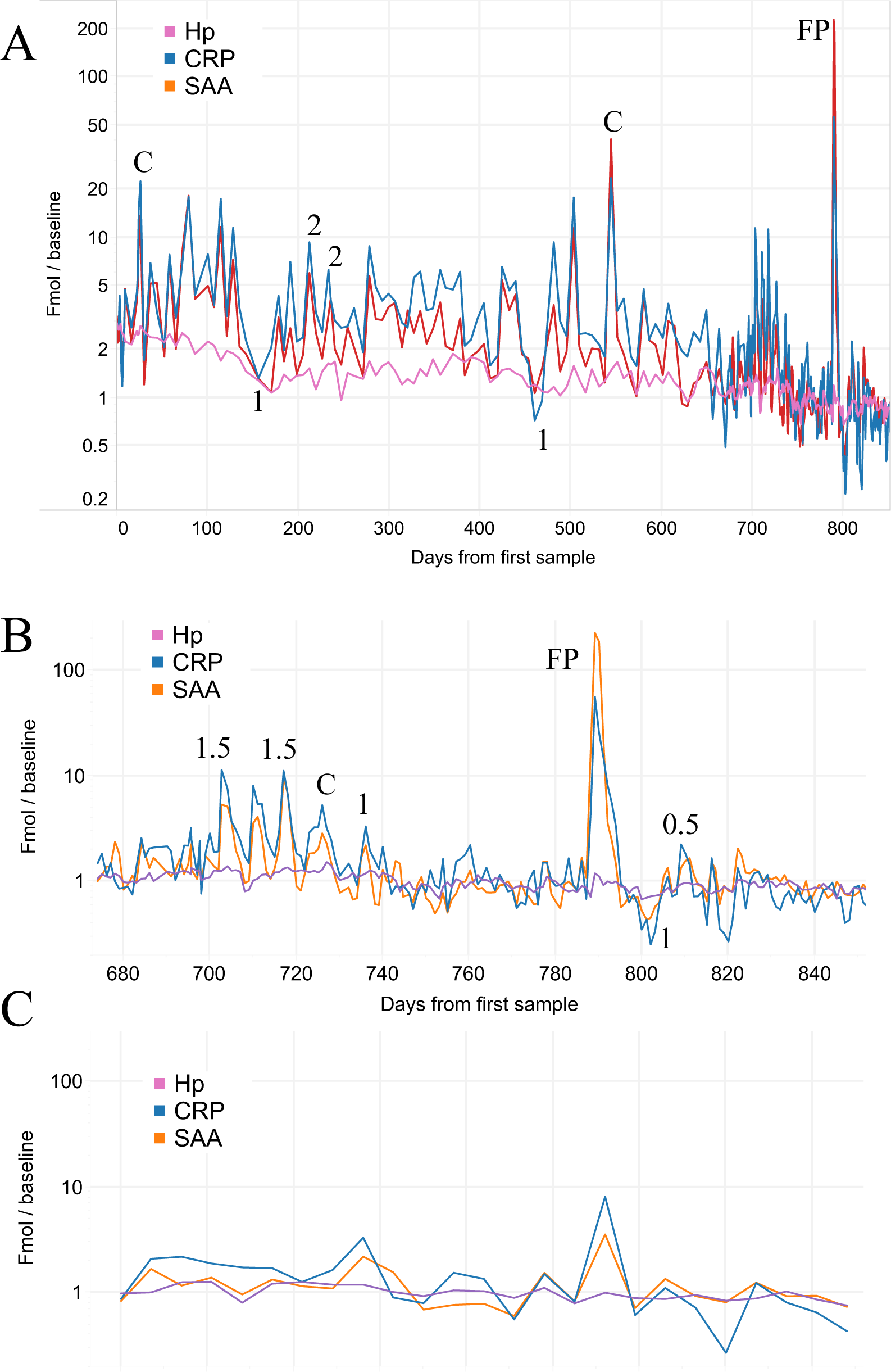
A) Amounts of Hp, CRP and SAA (normalized fmol divided by personal baseline median) in 284 samples from subject S-10 (Crohn’s disease) over 852 days (daily sampling from day 675, weekly sampling prior to that). Numbers indicate severity of Crohn’s attack (0-2 scale) assessed by subject, “ C” = cold, “ FP” = episode of food poisoning. B) Expanded view of daily samples. C) Weekly subsample (Thursdays) of daily sample data.

Fig. 3C shows results from weekly samples (Thursdays) extracted from the daily sample set shown in Fig. 3B. Weekly sampling gives a significantly different picture of inflammation activity than the daily samples in this case, consistent with the short timespan of most Crohn’s flares in subject S-10.

### Intense exercise

The most rapid APR protein changes observed were the result of intense physical training, in this case using dried venous blood samples from elite Brazilian beach volleyball athletes during a week of Olympic training. Four samples were collected on each training day: Figure 4 shows longitudinal changes in 4 teams (1-4) of two athletes (A and B) training together over the course of 4 training sessions and 2 subsequent recovery samples later in the week. Hp decreased significantly during training sessions, presumably due to complexation with Hb released through RBC hemolysis associated with violent muscle contraction [43], declining by a total of ∼10-60% among the 8 athletes by the end of session 4, and then recovering to normal levels 3 days later. Myeloperoxidase (MPO; reflecting the neutrophil count) increased by ∼10-100% during training sessions and rapidly returned to baseline [44], exhibiting large differences between athletes. CRP and SAA were not strongly affected during training sessions, though SAA appeared to decrease in most athletes during the 4 days of training. There were, however, several specific events indicating inflammatory responses from other sources: athlete A of team 3 (A3) began the week with CRP and SAA strongly elevated, and both declined according to typical half-lives. Athlete 4B experienced a large increase in neutrophils (MPO) during the first training session, followed by the largest observed CRP increase the following day, suggesting a potential injury (though none was noted by the coaches). While the overall pattern of Hp and MPO responses to intense training was consistent, individual athletes differed significantly in the magnitude of these changes.

**Figure 4:**
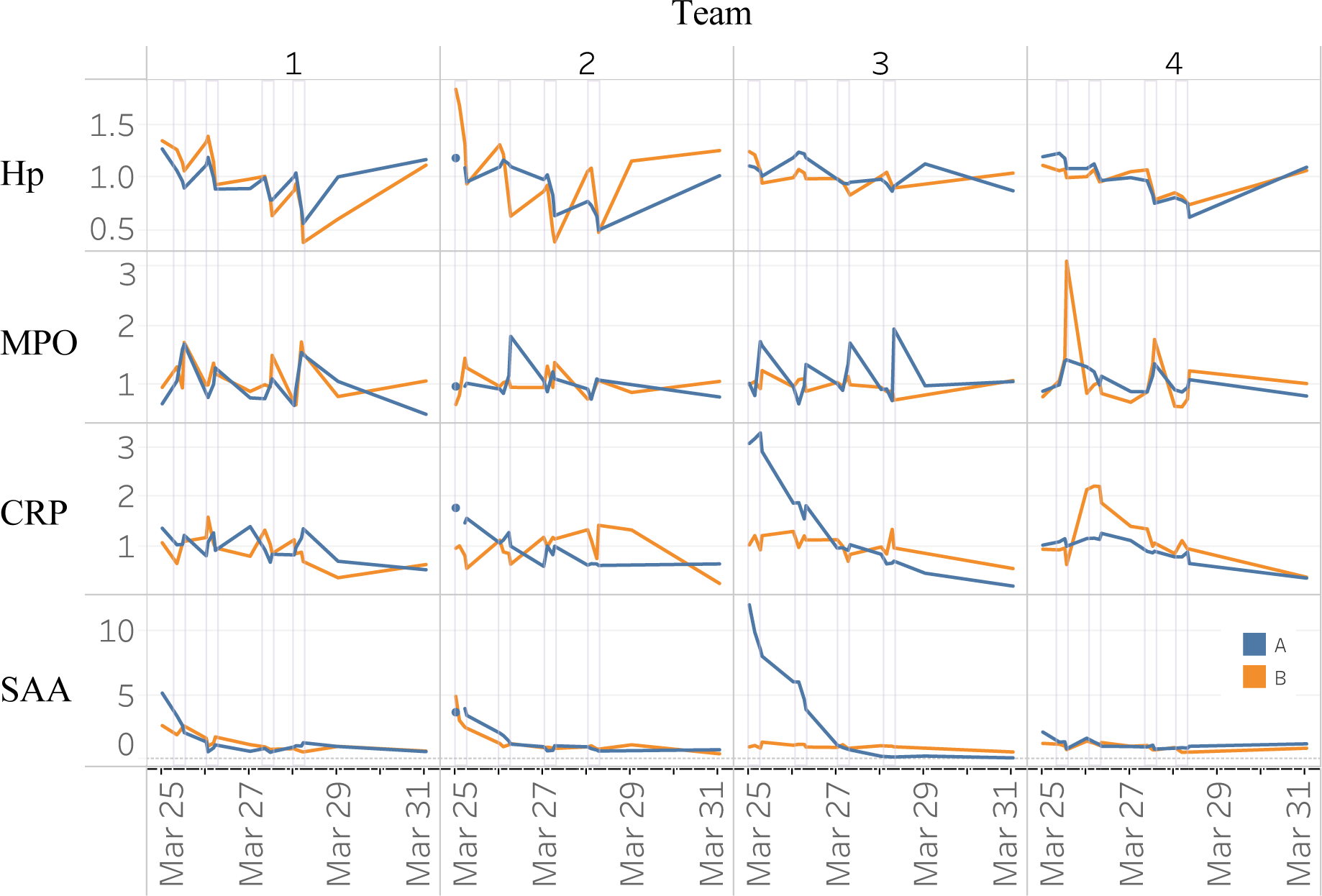
Amounts of Hp, MPO, CRP and SAA (normalized fmol divided by personal baseline median) in 8 elite beach volleyball athletes (4 teams of 2) during a week of Olympic training. Four training sessions are outlined in grey rectangles.

### Correlations among inflammation biomarkers across the data

Figure 5A shows a protein:protein correlation matrix calculated over all 1,522 samples from subjects in sample set I after volume normalization and division by personal baseline median values (to minimize the impact of between-subject differences in protein amount). The positive APR proteins showed significant correlations overall as expected, with the highest pairwise correlations occurring between proteins with similar time signatures (Figure 3E): thus CRP and SAA (both fast APR’s) show a pairwise correlation of 0.89; A1AG and Hp (both slow APR’s) correlate at 0.84. CRP and Hp (fast vs slow responses) showed a correlation of only 0.49. Alb and IgM showed strong negative correlations with the positive APR proteins, consistent with their known roles as negative APR markers. The 3 normalizing proteins Alb, Hx and IgM also exhibited strong negative correlation with each other, as expected given their collective balancing role in the volume normalization process.

**Figure 5:**
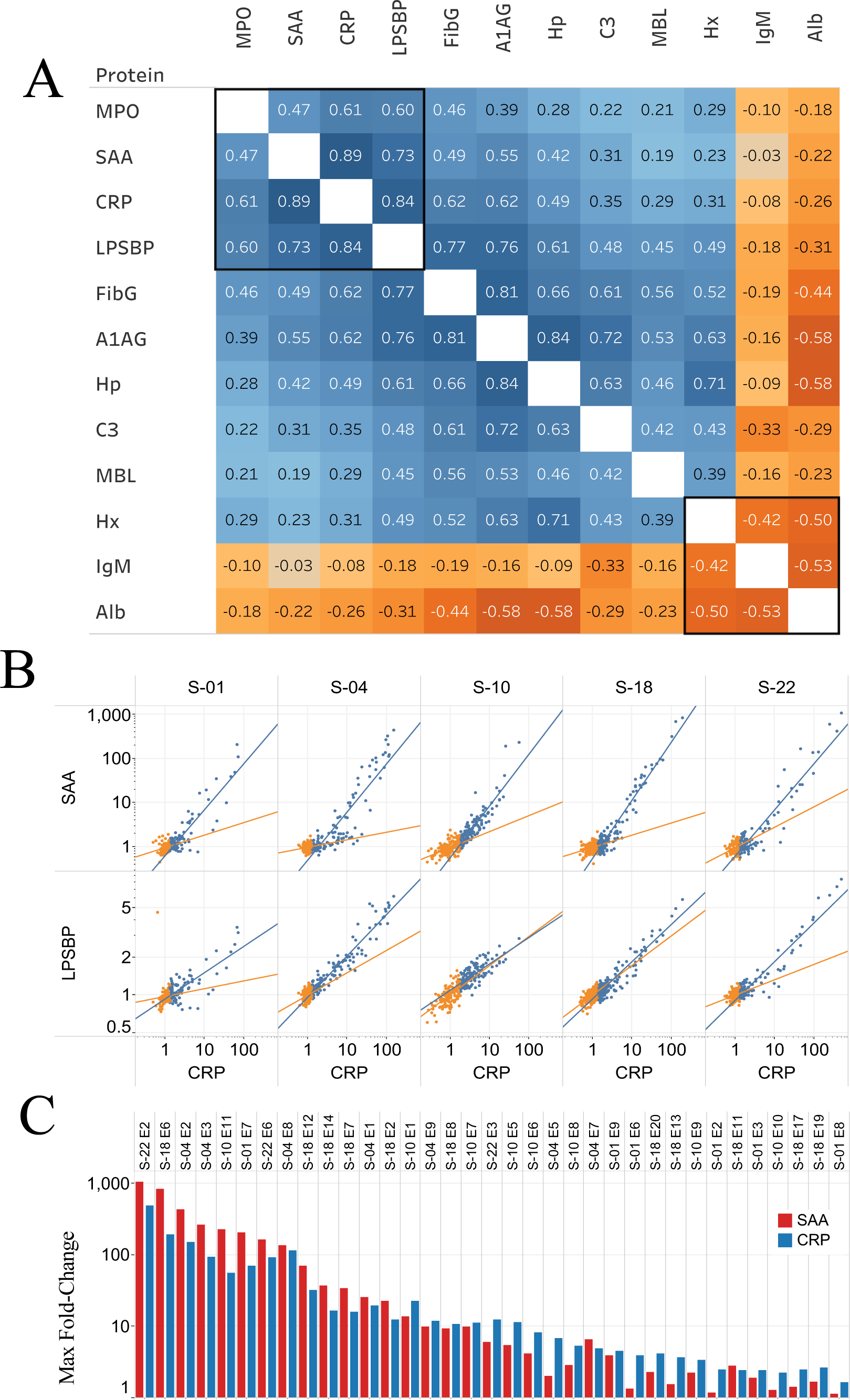
A) Protein:protein correlation matrix calculated over all samples from all subjects after sample volume normalization and division by personal baseline median values (to minimize the impact of subject:subject differences in protein amount). B) Scatterplots (normalized fmol divided by personal baseline median, log-log) relating CRP to SAA or LPSBP in the individual samples (dots) contributed by 5 subjects. Baseline samples (the half with lowest CRP) are shown in orange, and the higher half of samples in blue. C) Maximum fold-change from baseline in SAA and CRP during inflammation events.

### Non-linear regulation

Figure 5B explores the correlation of CRP with SAA and LPSBP in more detail using log:log plots in samples that are above (blue) and below (orange) personal median CRP levels for the 5 subjects with large sample numbers. During inflammatory events, where CRP varies by up to ∼100-fold, the SAA response was consistent across subjects, increasing by up to 1,000-fold over baseline in major infections. LPSBP levels increased in a similar pattern, but the observed increase was less than 10-fold at maximum. The strong relationships evident in the blue samples, while appearing linear in log:log format, are, in fact, non-linear (best-fit here by exponential relationships). This is confirmed in the set of events (Figure 5C) in which a local maximum SAA timepoint was determined (i.e., for which samples were available with lower levels just before and after a maximum), which shows that SAA’s maximum induction relative to baseline exceeds CRP’s for the highest-level events, but CRP generally achieves higher levels of induction than SAA for the smaller events. This non-linear relationship can be modeled by an exponential fit in which SAA is related to CRP raised to a power between 1.31 and 1.52, except for subject S-10 for which the power is 1.75.

Figure 5B also reveals significant correlations between CRP and SAA or LPSBP in the half of each subject’s samples (orange) with lowest CRP (i.e., in the baseline low-inflammation data). Such persistent correlations among proteins in what could be considered baseline samples demonstrate that small changes in APR proteins represent real biological signal (“ microinflammation”), not simply noise in pre-analytic and analytic steps.

### Visualization of inflammation responses and recovery

Differences in timing between earlier and later-peaking inflammation markers suggest an improved method for visualizing an inflammatory response trajectory as loops in two dimensions, starting at baseline, through injury and returning to baseline [45]. Figure 6A shows 3 such event trajectories in one subject (S-04) visualized by plotting SAA (fast response, up to 439-fold above baseline on a log scale) vs. Hp (slow response, maximum at 5.3-fold above baseline, also on a log scale) in successive DBS from the respective events. These events include a “ deep cough with earache” (S-10 E2; red), as well as the hip replacement surgery (S-10 E8; green) and “ nose cold” (S-10 E9; blue) events shown in Figure 2C. The largest magnitude event (cough) reaches maximum response at days eight and nine after first increase, while the much smaller response to a cold reaches maximum in three-four days. Response to the comparatively short two hr surgical intervention peaked in two-three days, and proceeded through initial healing (day eight), followed by a period of re-inflammation (days 10-27; coincident with increased use of pain medication) and recovery to baseline (day 86; blue).

**Figure 6:**
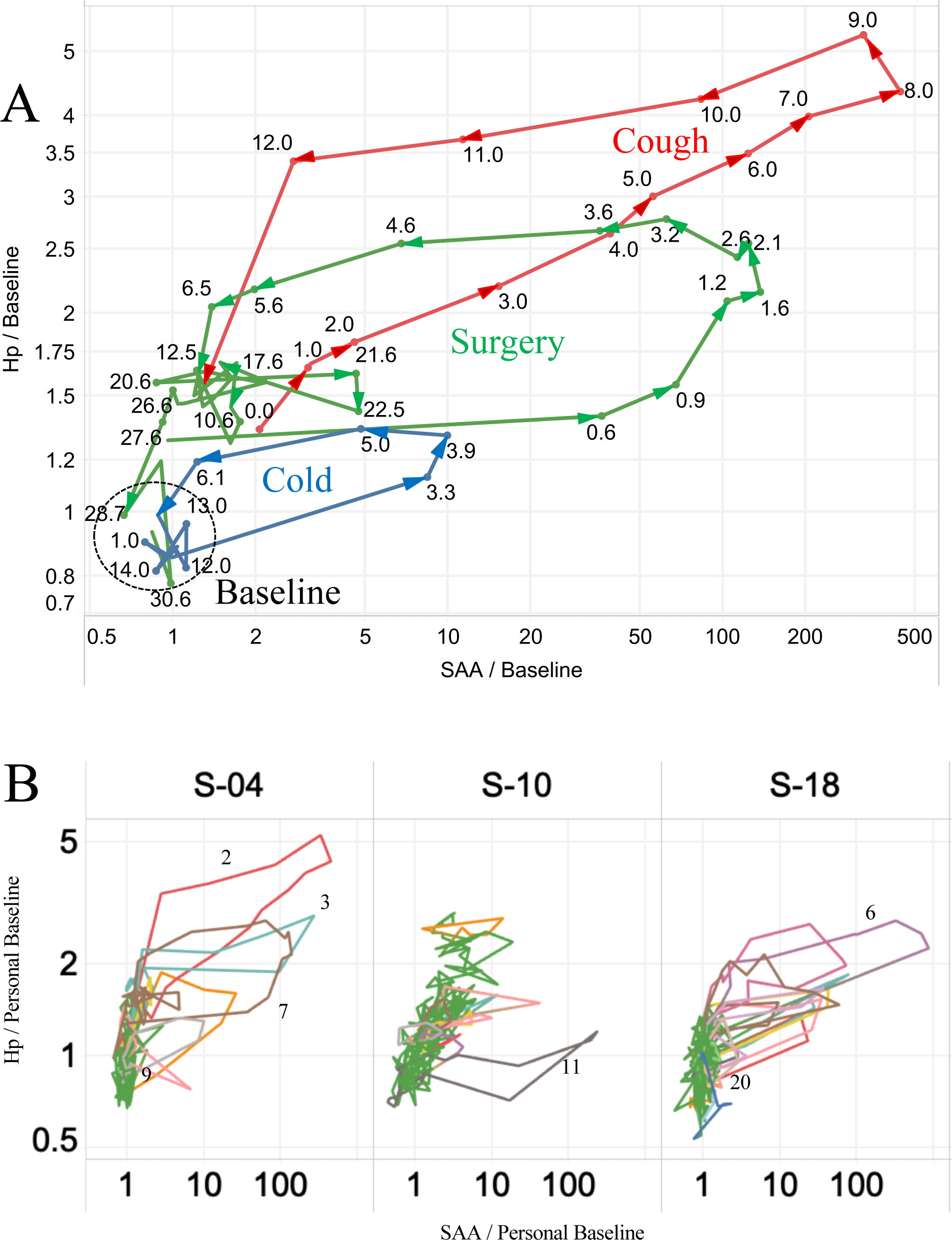
A) SAA vs Hp loop plot (normalized fmol divided by personal baseline median, log-log) for 3 inflammation events (Cold, Surgery, Cough) in subject S-04, with successive time points linked by lines to form loops. Numbers indicate days from beginning of inflammation event. B) Loop plots showing all samples for 3 subjects. Inflammation events color-coded, with baseline samples shown in green.

The same approach can be applied (Fig 6B) to visualize three subjects’ complete inflammation trajectories presented in Figure 1C. In these plots the identified inflammation events are drawn in colors, while the samples constituting baseline are shown in green. Subjects S-04 and S-18 each experienced a number of inflammation loops (all associated with infections except S-04 E7 (surgery). Subject S-10 (Crohn’s) has a different distribution of measurements, with only a single major infection (E11) and a wider range of Hp variation due to the progressive decline of this marker during the 2.5 yrs of sample collection.

## Discussion

Here we present a high-resolution multi-dimensional molecular picture of inflammation events and personal baseline biomarker levels in 16 individuals, combining dense longitudinal sampling *via* DBS, a broad panel of acute phase proteins and precise, multiplexed wide-dynamic-range quantitation using SISCAPA-MS. This picture captures details of inflammation processes on multiple time scales, reveals complex relationships among inflammation-related proteins and extends the measurable dynamic range downward to cover biology-driven fluctuations in APR baselines (“ microinflammation”). The results move beyond what can be observed through sporadic measurements of a single marker such as CRP and offer the potential for major improvements in diagnostic medicine and for monitoring subject responses in clinical trials.

While previous studies have examined many of these proteins alone, or occasionally in small groups, this study is unique in applying a broad set of APR markers across many indications at high frequency. Key to this broader coverage is selection of a group of APR proteins that vary widely in response kinetics, magnitudes and function. Each inflammation-associated protein biomarker shows a different characteristic time course of expression during an inflammation event. APR proteins, which are generally produced in the liver, exhibit a protein-specific early phase increase, likely governed by mRNA transcription and expression triggered by a short-lived cytokine signal. Among the APR proteins, LPSBP typically rises most rapidly, followed in order by SAA, CRP, A1AG, FibG, Hx, MBL, Hp, and C3. MPO, which in the context of whole blood serves as a surrogate for the neutrophil count, typically shows a spike just before or coincident with LPSBP, reflecting the fact that pre-existing neutrophils can be released from the bone marrow immediately upon cytokine signaling, without a time-lag required for new protein synthesis. APR proteins decline following an order (SAA, CRP, LPSBP, Hp, Hx, FibG, MBL, A1AG, C3) that is generally consistent with previously measured half-lives of CRP, LPSBP, A1AG and C3 of 26, 32, 104 and 84 hours respectively [46]. Proteins peaking earlier and later provide complementary information, analogous to the relationship between glucose and HbA1c measurements in diabetes: fast responders provide a measure of inflammatory stimuli in the previous 1-3 days, while the slower responders reflect stimuli that have occurred over much longer periods comprising a slower initial rise and a slower decay. Thus at any given timepoint, the combined set of proteins provides a readout reflecting inflammatory stimuli occurring over the previous 2-4 weeks

In addition to different expression times, the APR proteins also differ widely in the amplitude of changes during inflammation events. Proteins exhibiting fast responses typically show very large changes (10-to 1,000-fold from personal baseline values) while those showing smaller, though still statistically significant changes (in some cases 1.1-to 1.5-fold changes from personal baselines), typically show slow responses. Changes in Alb and IgM generally involve decreases from baseline during major infections (i.e., they are negative APRs, changing with polarity opposite to APRs like CRP).

The data also demonstrate directly, for the first time, that the quantitative relationships between strongly responding APR proteins such as CRP, SAA and LPSBP are non-linear; i.e., producing different relative change magnitudes in large *vs* small scale inflammation events. The APR panel used here thus achieves its overall design goal: to include proteins responding differently in time, magnitude, polarity and functional responses to inflammatory signals.

Distilling these complex data in a way that provides maximum value in clinical diagnostics or pharmaceutical trials presents several challenges. In the past, efforts to move beyond single point CRP measurements have explored two scenarios: a multi-protein ratio at one time point, or alternatively a single protein measured at multiple time points. Both approaches have significant limitations.

In the first scenario, several ratios involving CRP have been explored for clinical diagnostic applications, including, for example, the ratio of SAA to CRP, proposed as an indicator of infectious disease severity in children [47]. In our results this ratio does appear to distinguish between large and small infection responses when measured at the peak of response (Fig 5C); however, the ratio changes so rapidly during an infection (up to 20-fold change over the course of a few days as shown in Fig 2B) that sampling at any point off-peak can generate an incorrect prediction. Likewise the ratio of fibrinogen to CRP has generated equivocal results [48–50] since it also varies in time due to the difference in expression time courses for the two proteins. Somewhat more success has been achieved using the ratio of CRP to albumin as a long-term outcome predictor in pancreatic cancer [51], surgery in the elderly [52], sepsis [53], and stent restenosis [54], while a similar 3-level index combining CRP and Alb measurements (the Glasgow Prognostic Score [55]; GPS) has achieved more widespread use as an outcome predictor in many cancers. The greater success of the CRP/Alb ratio and GPS may be attributed to the fact that the components move in opposite directions during the APR (being positive and negative APR respectively), so that inflammation-driven change of either protein from baseline drives the ratio upwards. Nevertheless, the fact that individual APR proteins follow different time courses, making any ratio between proteins time-varying, substantially limits the utility of protein:protein ratios at any single timepoint.

The second scenario, using an index of CRP changes over time, has been more generally successful for evaluating infection severity [56], risk of death in intensive care [57], presence of post-surgical infection [58] and likelihood of success or failure of treatment in community-acquired pneumonia [59], sepsis [60], Crohn’s disease [61] and stem cell transplantation [62]. These methods make use of changes occurring between sample timepoints and thus provide more information than single point CRP measurements. Nevertheless, because of the fast-response nature of CRP, they do not incorporate information on inflammation levels outside narrow time-windows bracketing sample collection, nor, without effective prior measurement of personal baseline levels, can they provide a personalized indication of inflammation severity.

A more powerful approach explored here incorporates measurements of multiple proteins at multiple time points. As a first step, we have used two APR proteins responding on different timescales (fast-responding SAA and slower-responding Hp) which allows inflammation events to be visualized in two dimensions as shown in Fig. 6A. In this representation, inflammatory events unfold as counterclockwise loops [45], beginning at baseline values for both proteins (coordinates near 1:1 in the lower left), moving first to the right (rise of fast APR), then up (rise of slow APR), then left (decline of fast APR) and finally down towards baseline (decline of slow APR). Loop size and distance from the subject’s baseline samples indicate the magnitude of the response: the 2-week long upper respiratory infection (“ Cough”) extends further than the surgical loop, which in turn is larger than the loop corresponding to a cold. Deviations from a smooth loop trajectory are attributable in the surgical case to post-surgical strains noted by the subject and correlated to increased pain medication (Fig 2C and 2D). Any two adjacent (i.e., consecutive) samples on such a loop generate a directional line segment whose position and slope indicate a) the size of the loop (whose area approximates the intensity and duration of the whole response) and b) where the subject is on the loop trajectory at that time (i.e., position on the path from insult, through recovery and back to baseline). Given the relatively smooth structure of these loops, it appears that comparison of two samples taken 6 hr (0.25 d) apart would provide a useful estimate of a subject’s position and direction on the response loop, given the assay precision and baseline data obtained here.

Our long-term objective, however, is to create a true multi-dimensional, personalized longitudinal inflammation model including additional APR components and related features such as neutrophil bursts (measured here as released MPO) preceding APR and adaptive immune responses (measured here as IgM) following some infections. Such a model will incorporate the temporal and magnitude relationships among the biomarkers (including the non-linear regulatory effects first reported here), fitting multi-protein data from observed events to generate important derived parameters. These include the integrated time and intensity of inflammation (the loop areas in Fig. 6A being analogous to area under the curve for drug effects), as well as inferred time of the start of inflammation (when sampling only begins after an event has started) and deviation from a classic response trajectory (as a means for distinguishing inflammation events with different causes). Some of the required methods may be adapted from classical pharmacodynamic (PD) modeling, in which biomarkers are used to measure response to a known cause (e.g., a pharmacokinetic (PK) drug dosing model). Simple PD models using CRP as a single biomarker have been developed and used successfully in clinical trials of antibiotics [26–28], various biologics [20,21,23,24]; antisense to CRP [25], and a MAPK inhibitor [33]. However PD-like models of inflammation have not been adopted in clinical diagnostics despite a small handful of successful examples [63], due in part to the rarity of applicable longitudinal samples and in part to the inverse nature of the problem: whereas in PK/PD modeling the goal is to model the effects of a known cause, in diagnostics one seeks to infer the cause from the effects on biomarkers. Personalized PD inflammation models, developed using baselines and responses to inflammation events (e.g., vaccinations, colds) in a subject’s longitudinal samples, would provide increased precision in tracking inflammatory phenomena and optimal design of sampling protocols. Additional methods for uncovering disease:biomarker relationships, including machine learning approaches developed in “ big data” applications, are also likely to benefit from personalized longitudinal multiplex data described here.

A number of novel observations emerged from examination of our extended longitudinal sample series. First is the unexpectedly large number of identifiable inflammation events, most of which were apparently due to infections in apparently normal subjects. A surprisingly high proportion (12-45%) of each subject’s samples showed indications of inflammation, whether defined as CRP levels >2-fold above personal baselines, or inclusion in the 57 discernable APR events. Similar proportions are obtained when considering only those samples collected on a near-daily basis (about 1/3 of the samples), in which the timing of collection should be less susceptible to subjects’ selection bias. Such high event frequencies, if confirmed in a larger subject population, suggest that unrecognized short-term “ sub-clinical” inflammatory events are likely to occur frequently during investigational studies and drug trials, and perhaps at even higher frequency in unwell individuals (of which the Crohn’s disease subject is an example). Such changes may impact behavior of drugs not directly related to inflammation, e.g., due to the well-known binding of many small molecules to the APR proteins albumin and A1AG [64].

The frequency of small inflammation events also suggests that randomly-timed single CRP measurements (such as those included in annual checkups) may overestimate a patient’s true baseline level if collected during an inflammatory episode (i.e., 12-45% of the time according to our estimate of samples indicating inflammation), and thus bias predictions of cardiovascular disease risk [3,65]. Conversely a single CRP measurement is likely to underestimate the likelihood that a patient has inflammatory episodes when used as a qualification for anti-inflammatory therapy [66]. A clear improvement will be to use personal median baseline APR levels (including CRP) derived from a series of longitudinal samples thus providing a significantly better risk estimate by capturing baseline inflammation separately from fluctuations related to transitory subclinical events.

Characterizing personal baseline inflammation proved to be more challenging (and interesting) than anticipated. In the half of each individual’s samples with the lowest CRP (presumably the samples showing the lowest levels of inflammation) we observed persistent correlations between CRP, SAA and LPSBP, indicating that a significant amount of the observed baseline variation represents low-level, but real, biological response fluctuations (“ microinflammation”), rather than analytical “ noise”. As a result, the spectrum of inflammatory responses measurable using APR biomarkers appears to extend smoothly from +/- 10% (i.e., 0.9-1.1-fold) to 1,000-fold, a total dynamic range of over 10,000. This observation suggests the possibility of correlating detectable subclinical inflammatory triggers with a variety of contextual data including diet, environmental exposures, recurring infections and chronic inflammatory disease.

The 57 discrete inflammation events we cataloged included events driven by a variety of causes, both “ planned” (surgery, vaccination, and intense exercise) and “ unplanned” (infections, and episodic inflammation related to Crohn’s disease). Given the small number of subjects covered, these observations represent individual case reports and require follow-up in well-designed studies of larger cohorts to confirm the generality of our observations.

Total hip replacement surgery in subject S-04 represented a “ planned” inflammatory intervention, whose known start time and 2 hr duration allowed precise timing of samples with respect to the inflammatory stimulus (Figure 2C, D, E and Fig. 6A). In this case SAA rose rapidly to maximum in less than 48 hours and fell to near baseline by day seven, while Hp and other slow APR proteins reached a maximum on day four and did not return to baseline for four-eight weeks. During the post-surgical recovery period, three clearly discernable peaks (∼9, 18 and 22 days after surgery) interrupted the recovery trajectory, the latter two of which correlate in time with physical “ strains” noted by the patient, causing increased requirement for pain medication (Fig. 2D). Between 50-60 days post-surgery, the levels of APR proteins reached a new baseline significantly below pre-surgery levels, a decrease in inflammation that provides a quantitative measure of the benefit of replacing an arthritic hip joint. The level of temporal detail observable by this approach provides a much more complete picture of recovery, including interruptions, than shown in previous studies using a single marker (typically CRP) to detect post-surgical infection [58] or to assess surgical damage [67].

A series of infection events generated the largest APR protein changes observed here, seven of which resulted in SAA levels ≥100-fold above personal baselines (Fig. 2A). These events, and a series of smaller events usually attributed to common “ nose colds”, provides evidence of a non-linear relationship between maximal CRP and SAA inductions, as well as the lack of synchrony mentioned above which limits use of the SAA/CRP ratio. Influenza vaccination resulted in detectable APR responses that were ∼100-fold lower than those observed with the major infections. In addition to acute phase proteins, we also observed increases in IgM and MPO in a subset of infections. While most infections did not cause measurable increases in IgM, three episodes resulted in increases of 30-50% in total IgM 8-10 days after SAA peaked (Supplementary Fig. 4), after which the levels subsided to pre-existing baseline levels. The production of such large amounts of IgM after specific infection events is consistent with the expected timeframe for an adaptive immune response to a pathogen, and may provide an opportunity to identify endogenous human monoclonal antibodies from DBS samples with potential therapeutic value as anti-infectives. In contrast, MPO, which serves in whole blood as a surrogate for neutrophil count, was frequently increased at the earliest stage of infection followed by spikes every two or three days (Supplementary Fig. 3), consistent with periodic release from and regeneration of neutrophil pools in the bone marrow [68].

In a single case of Crohn’s disease we observed a higher frequency of APR events than in the other subjects, including 8 subject-identified “ Crohn’s attacks”, 3 “ colds” and one instance of food poisoning. Interestingly, 3 of the Crohn’s attacks occurred at times of minimal APR levels, while the others coincided with APR spikes of 2-to 10-fold above baseline levels. The brief timecourse of the attacks highlights the necessity for frequent sampling as a basis for selecting an optimal sampling strategy for any given condition: a weekly sampling schedule would have completely missed most of the events (Fig 3B and C). A significant number of similar intensity APR increases were not annotated and may represent subclinical events in unknown cause. Of practical importance, over a 2-5 yr period, average CRP, SAA and Hp levels decreased alongside steady improvement in the subject’s control of the disease. Since the relationship of Crohn’s disease activity to inflammatory biomarkers such as CRP may be useful in a “ treat-to-target” approach to biologic therapy [69], it will be important to further elucidate these relationships in multiple subjects.

Intense exercise with accompanying heat stress and strong sun, in this case occurring during Olympic training for beach volleyball, was included here to assess the potential magnitude of very short-term physical effects on APR responses. The most significant changes observed were large decreases in Hp (presumably due to rapid removal of Hp:Hb complexes formed as a result of exercise hemolysis [70]) and increases in MPO (presumably due to release of pre-formed bone marrow neutrophils) during daily training sessions, neither of which depend on expression of new protein. One substantial induction of CRP was noted in athlete 4B, occurring ∼18 hr after the largest spike in MPO, possibly indicating minor muscle damage.

Taken together, these results demonstrate that DBS collected under field conditions and stored for periods up to nine years can be analyzed by SISCAPA-MRM to deliver high-precision biomarker measurements spanning a very wide dynamic range. The automated workflow used here [39], together with a personalized normalization strategy to remove DBS volume variation, reduced assay CVs to ∼5% in standard samples, a level substantially below previously estimated within-person variation [71].

We note several limitations of the present study. While the sample numbers analyzed here exceed those in all but a handful of biomarker studies, the number of subjects is modest, with only 5 individuals contributing more than 100 longitudinal samples each. Future studies will require larger numbers of subjects in order to generalize conclusions about the frequency, scale and time courses of inflammation events, to link this data with specific causes and outcomes, and to determine optimal sampling frequencies for specific applications. While not strictly necessary in studies such as ours that refer biomarkers to personal baselines, we also look forward to reporting calibration of the individual biomarkers against validated clinical assays, and rigorous comparison of dried capillary blood with liquid venous serum (manuscripts in preparation).

### Future Perspective

In the future, a number of important opportunities can be addressed using inflammation and other protein panels measured in subject-collected DBS and interpreted against carefully defined personal baselines. Aside from obvious diagnostic needs in infectious and chronic inflammatory diseases, APR responses are frequently noted in cancer biomarker discovery studies and are likely to prove useful in monitoring cancer patients during treatment and in detection of recurrence. Clinical trials of anti-inflammatory biologics can likewise benefit from detailed tracking of APR responses covering a wider range of timescales than can be obtained with CRP alone. In addition, patients can potentially benefit from more precise monitoring of inflammation in relation to lifestyle changes, medical challenges and associated treatments.

### Executive Summary

#### Background

- Inflammation is a key clinical feature of many disease states and its reduction is the goal of a rapidly expanding range of biologics and small molecules
- C-reactive protein is currently used as a single diagnostic indicator of inflammation in patients and is the primary biomarker of efficacy in anti-inflammatory drug trials
- Here we explore a higher-resolution picture of inflammation, using multiple protein biomarkers and high-frequency sample collection to provide deeper insights into structure of inflammatory events as well as asymptomatic normal baseline.

#### Experimental

- A unique set of 1,658 dried blood samples (DBS) was collected from 16 individuals over periods up to 9 years, including daily collection for extended periods
- A panel of nine positive and three negative APR proteins was measured by SISCAPA-LC-MRM mass spectrometry
- Spot-to-spot volume variation in dried blood spots (DBS) was reduced substantially by normalization of a three-protein panel, providing total workflow assay CVs on replicate DBS samples of 2.5-6%

#### Results

- A total of 57 discrete inflammation events were observed related to major infections, vaccination, surgery, extreme exercise and Crohn’s disease
- APR proteins were differentially regulated during these events, with unique non-synchronous time courses and non-linear regulatory relationships that require PD-like modeling and interpretation more advanced than currently-used ratios
- CRP, SAA and LPSBP remained correlated in the subset of samples with lowest CRP, indicating that very low level “ microinflammation” phenomena can be measured and extending the dynamic range of measurable APR to >10,000.

#### Discussion

- Multiplexed measurement of a broad panel of acute phase response (APR) proteins, including CRP, at frequent timepoints using DBS microsamples provides improved insights into the magnitude and dynamics of inflammation processes
- The approach provides personal baselines for each protein, allowing improved diagnostic performance
- Clinical trials of anti-inflammatory biologics, antibiotics, and cancer therapeutics can benefit from improved multi-marker pharmacodynamic models of inflammation.

## Materials and Methods

### Samples

A unique set of 1,522 long-term longitudinal capillary blood DBS samples (sample set I) was self-collected at home between 2008 and 2017 by 8 participants using lancet finger-pricks (Medlance Plus Extra or Special, HTL Strefa; Medline, Cat. No. HTD7045BX) and dried on Whatman 903 Protein Saver DBS cards (Table 1). The DBS cards were stored at 4° C in the presence of desiccant except for brief periods at room temperature or at −20° C and were barcoded prior to analysis. Transportation of specimens to the laboratory for processing followed the guidelines provided by the US Center for Disease Control and Prevention (CDC) for shipment of DBS specimens. In addition, a set of 140 EDTA-treated venous whole blood samples (sample set II) was collected from eight Brazilian professional athletes during four consecutive days of Olympic-level beach volleyball training in a study organized by Prof. L.C. Cameron and A. Bassini-Cameron (Universidade Federal do Estado do Rio de Janeiro, Brazil). Samples were collected before breakfast, before training, after training and 60 min after training) and after 1 or 3 days of recovery, and were spotted and dried on Whatman 903 filter paper.

### SISCAPA-LC-MRM protein measurement

A panel of proteins of known clinical significance was measured using SISCAPA-MRM mass spectrometry. These included a broad set of APR proteins and other inflammation-related proteins reported here (Table 2) and other biomarkers to be reported elsewhere. Sample preparation and SISCAPA peptide enrichment were performed using an automated protocol essentially as described [37–39,72] for all three datasets, with different, but largely overlapping, sets of measured protein targets. Target protein amounts were expressed as femtomole (fmol) of proteotypic peptide in each sample, calculated by multiplying the observed peak-area ratio (PAR; the ratio of the endogenous target peptide MRM peak area to that of the stable isotope standard (SIS)) by the known amount of added SIS. SIS peptides were labeled with 13C, 15N lysine or arginine as the c-terminal amino acid.

### Data analysis

PAR data were assembled in Tableau Prep Builder (www.tableau.com) and joined with Excel tables defining sample characteristics (e.g., date of collection, subject contextual health notes, SIS concentrations, analytical run structures, etc). Data analysis and visualization were performed in Tableau Desktop (Tableau Software, Inc.).

To minimize the effect of variations in plasma volume between DBS from the same individual (typically +/- 10-15%), we refined and applied a method developed previously [39] to normalize plasma volume using a sample-specific scale factor computed from an equally-weighted combination of three proteins (albumin, hemopexin and IgM) whose abundances are usually very stable within individuals over time.

Replicate standard DBS samples were included in each 96 well plate processed for SISCAPA measurement, normalized using the above methods and used to evaluate the precision of each assay in each dataset. The average coefficient of variation (CV) across the assays used in Datasets A, B and C were 6.7%, 5.9% and 4.5% respectively, consistent with expectations for clinically useful assays.

After this subject-specific normalization, subject-specific baselines were computed for each protein as the median level in the half of samples with lowest CRP (i.e., the samples showing least evidence of current inflammation). Fold-changes were computed relative to these baselines and so focus on relative responses in the individual rather than absolute scale of change.

## Data Availability

The data will be made available to collaborators by agreement with SISCAPA Assay Technologies.

## Supplementary data

To view the supplementary data that accompany this paper please visit the journal website.

## Ethical conduct of research

The authors state that they have obtained appropriate institutional review board approval or have followed the principles outlined in the Declaration of Helsinki for all human or animal experimental investigations. In addition, for investigations involving human subjects, informed consent has been obtained from the participants involved.

## Acknowledgments

The authors thank all sample donors for their participation, and Jordan Kallman and Steven Skates for helpful, insightful discussions..

## Author contributions

NLA and MR designed the project with input from MP and TWP; LCC and ABC designed the Sportomics study, collected samples from athletes and reviewed the manuscript; MR and RY carried out analytical workflow; NLA carried out data analysis in Tableau and wrote a draft manuscript which was edited by MR, MP, LCC and TWP.

## Supplementary information for

### Details of datasets

Dataset A results have been previously described in part [39] and were obtained using a liquid chromatography tandem mass spectrometry (LC-MS/MS) platform consisting of an Agilent 1290 LC system operating at 600 μL/min and an Agilent 6490 triple quadrupole MS. Replicate standards were prepared as punches of DBS (4 punches of 1.5 mm diameter filter paper were used per well). Datasets B and C employed an Eksigent microflow LC system operating at 10 μL/min connected to a Sciex 6500 Q-trap MS. Dataset B (436 samples from four subjects) focused on measurements of a panel of gastrointestinal inflammation related proteins. Replicate standards were prepared by pipetting whole blood onto pre-cut 6 mm diameter disks of Whatman 903 filter paper in sample wells. Dataset C was comprised of two sequential panels applied to the same DBS digests.

Nine peptides were measured in all datasets (including Alb, Hx, and IgM) and 12 were measured in both Datasets A, C and D. As part of our continuing efforts to increase assay precision, the 27-plex used in Dataset C was measured as two sequential SISCAPA multiplex subpanels, both of which included the normalizing proteins albumin (Alb), hemopexin (Hx) and total immunoglobulin M (IgM). Datasets A and B were obtained using a single MRM per peptide (measured as a single multiplex panel), while Dataset C used the average of 1 to 3 of the best quality MRM transitions available for the respective peptides. Datasets B and C were each merged with Dataset A using a run-to-run scale factor for each protein in common between datasets in order to account for potential shifts in the absolute amounts of the SIS peptides during storage between the analytical runs. The scale factor was derived from the ratio of average values for a protein in subject samples that were analyzed in both Datasets (88 samples analyzed in both Datasets A and B, and 19 samples analyzed in both Datasets B and C, in each case using duplicate punches from the same DBS card). When samples from the same subject were shared between datasets this ratio was subject-specific; for subjects without shared samples an average of the subject-specific ratios was used.

PAR measurements in Datasets A and D were determined using MassHunter Quantitative Analysis (v. B.05.02), while in Datasets B and C the Sciex MultiQuant program (v. 3.0.2) was used.

CVs were typically somewhat higher in Datasets A and B, which were analyzed earlier in the process of assay optimization. In Dataset C the CV over all 26 peptides measured in two successive multiplex enrichments from 4 replicate dried blood standards in each of nine 96-well plates (72 measurements) was reduced from 10.2% prior to volume normalization to 4.6% after normalization (Supplementary Table 1). Similarly, the overall average CV in Dataset A was reduced from 9.4% to 7.4% by normalization. In Dataset B, normalization had little effect (6.5% vs 6.1%) because in this case the standards were prepared by drying the same measured volume of whole blood onto pre-cut filter paper disks in wells, such that there was little or no volume variation to correct. In Dataset C, key inflammation markers such as C-reactive protein (CRP), serum amyloid A (SAA) and lipopolysaccharide binding protein (LPSBP) showed normalized CVs in the standard samples of 3.3% - 3.7%.

**Supplementary Table 1.**

| Protein | Dataset A Standards |  | Dataset B Standards |  | Dataset C Standards |  |
| --- | --- | --- | --- | --- | --- | --- |
|  | CV | CV | CV | CV | CV | CV |
|  | Before Vol | After Vol | Before Vol | After Vol | Before Vol | After Vol |
|  | Normalization | Normalization | Normalization | Normalization | Normalization | Normalization |
| A1AG | 11.3% | 8.6% |  |  | 9.2% | 4.7% |
| Alb | 7.2% | 1.9% | 7.2% | 5.7% | 10.4% | 1.9% |
| C3 | 6.2% | 2.3% |  |  | 9.3% | 5.5% |
| CRP | 14.1% | 13.1% | 6.8% | 6.8% | 9.8% | 3.3% |
| FibG | 6.9% | 5.3% |  |  | 10.6% | 4.3% |
| Hp | 10.2% | 9.4% | 5.3% | 5.5% | 6.8% | 9.4% |
| Hx | 6.4% | 1.4% | 4.6% | 3.0% | 10.0% | 1.5% |
| IgM | 6.1% | 2.6% | 6.1% | 4.6% | 9.8% | 2.1% |
| LPSBP | 7.6% | 6.5% | 6.4% | 6.2% | 10.2% | 3.7% |
| MBL | 8.1% | 7.8% | 11.1% | 11.2% | 14.2% | 12.1% |
| MPO | 11.7% | 13.1% | 5.8% | 5.5% | 10.5% | 9.7% |
| SAA | 10.9% | 8.8% | 5.0% | 4.8% | 11.6% | 3.7% |
| Average | 8.9% | 6.7% | 6.5% | 5.9% | 10.2% | 4.5% |

**Supplementary Table 2.**

| Subject | Total samples | Inflammation events | Samples with CRP>2-fold over baseline | Samples in events | Fraction samples with CRP>2-fold over baseline | Fraction samples in events |
| --- | --- | --- | --- | --- | --- | --- |
| S-01 | 186 | 9 | 47 | 91 | 25% | 49% |
| S-04 | 250 | 9 | 78 | 109 | 31% | 44% |
| S-07 | 49 |  | 6 | 0 | 12% | 0% |
| S-10 | 284 | 11 | 128 | 67 | 45% | 24% |
| S-17 | 87 | 1 | 30 | 8 | 34% | 9% |
| S-18 | 411 | 20 | 77 | 133 | 19% | 32% |
| S-20 | 45 | 2 | 14 | 8 | 31% | 18% |
| S-22 | 210 | 6 | 45 | 34 | 21% | 16% |

**Supplementary Fig 1.**
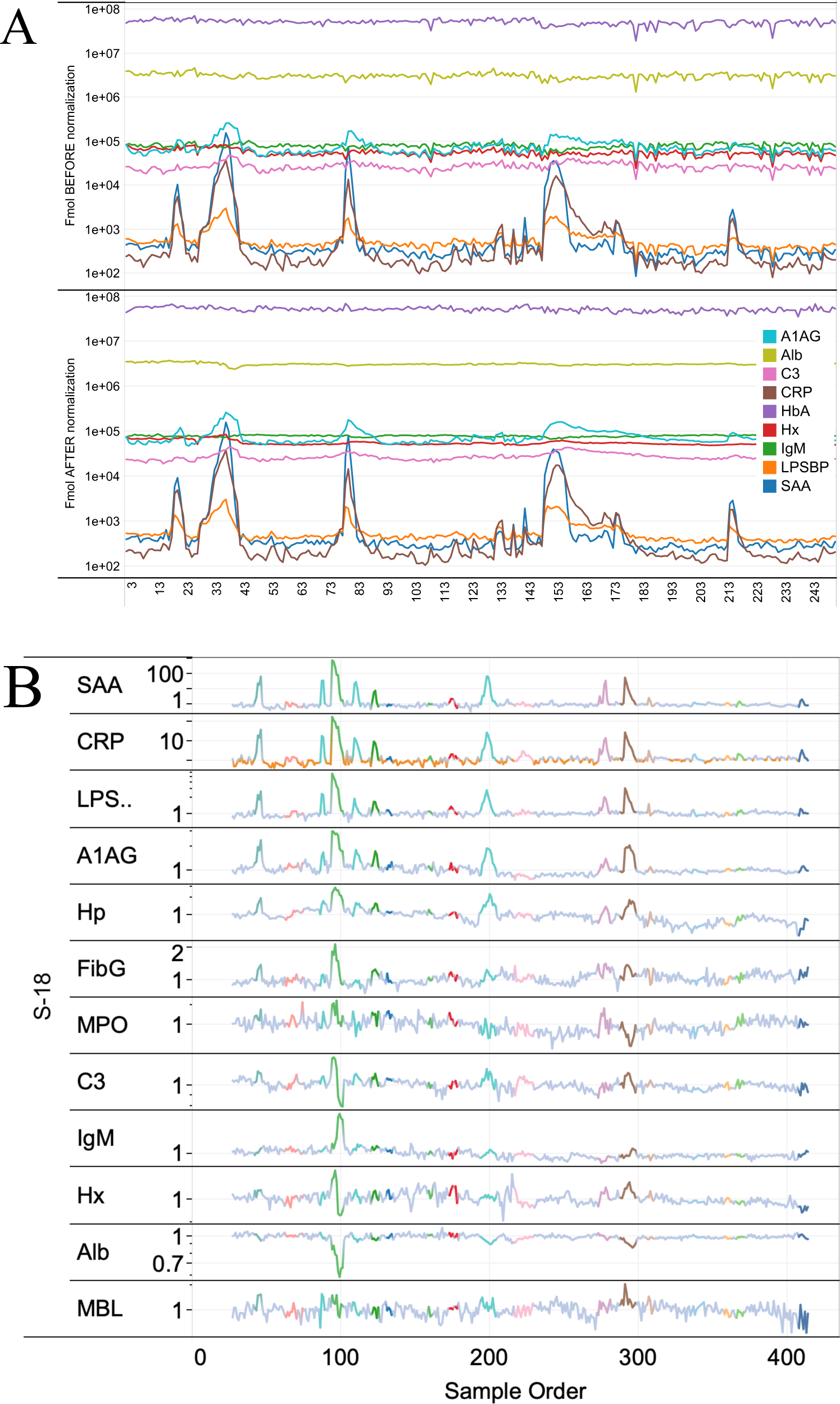
A) Amounts (fmol of peptide) for 9 proteins measured in 255 longitudinal DBS samples from subject S-04 before (top panel) or after (lower panel) DBS volume normalization by Alb, Hx, IgM. B) Amounts of 12 proteins mrasured in 411 serial samples from subject S-18 after normalization and division by personal baseline values.

**Supplementary Fig 2.**
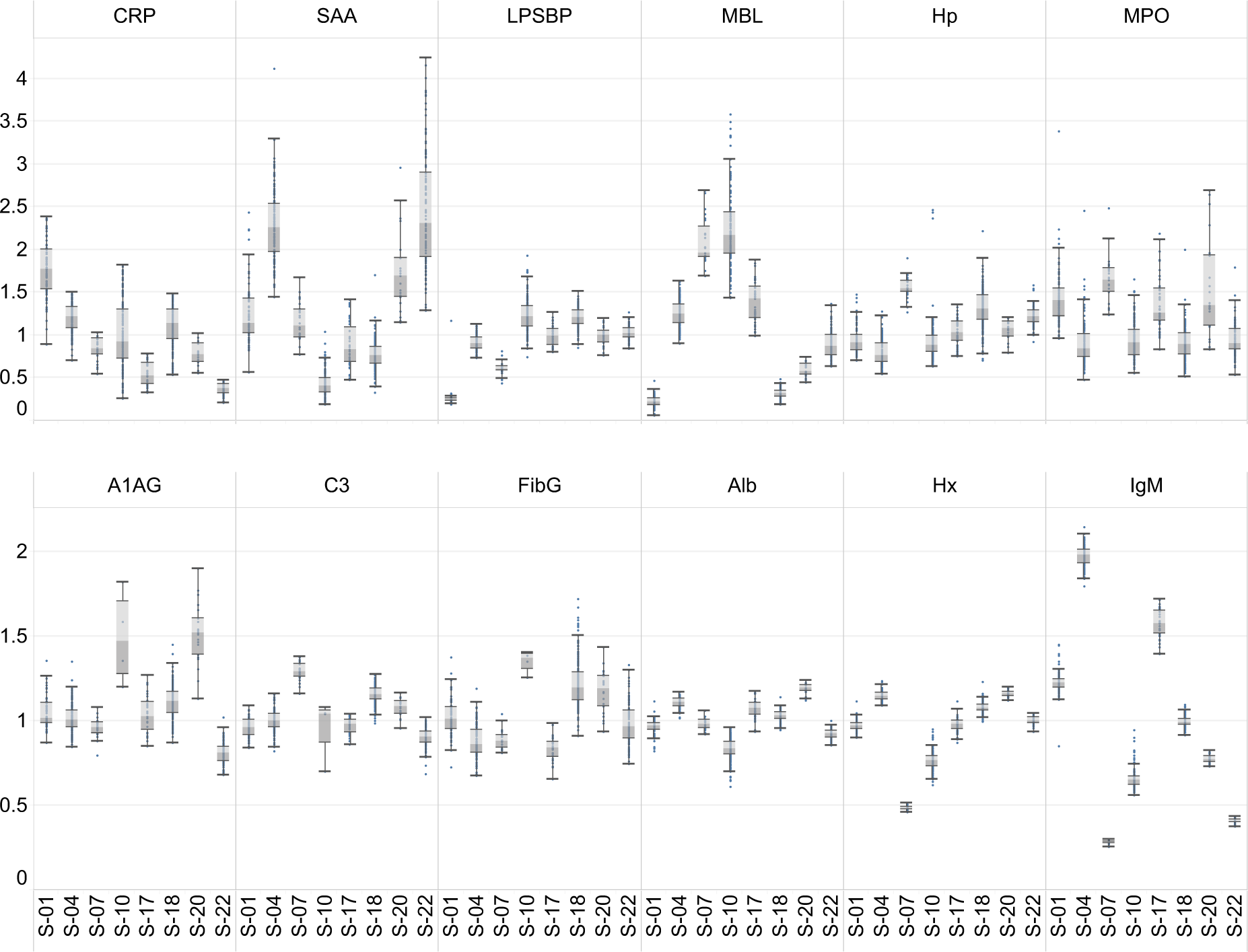
Whiskerplots of the amounts of 12 proteins in the baseline samples of each of 8 subjects. Protein amount has been normalized between proteins by dividing by the median amount in all subjects.

**Supplementary Fig 3.**
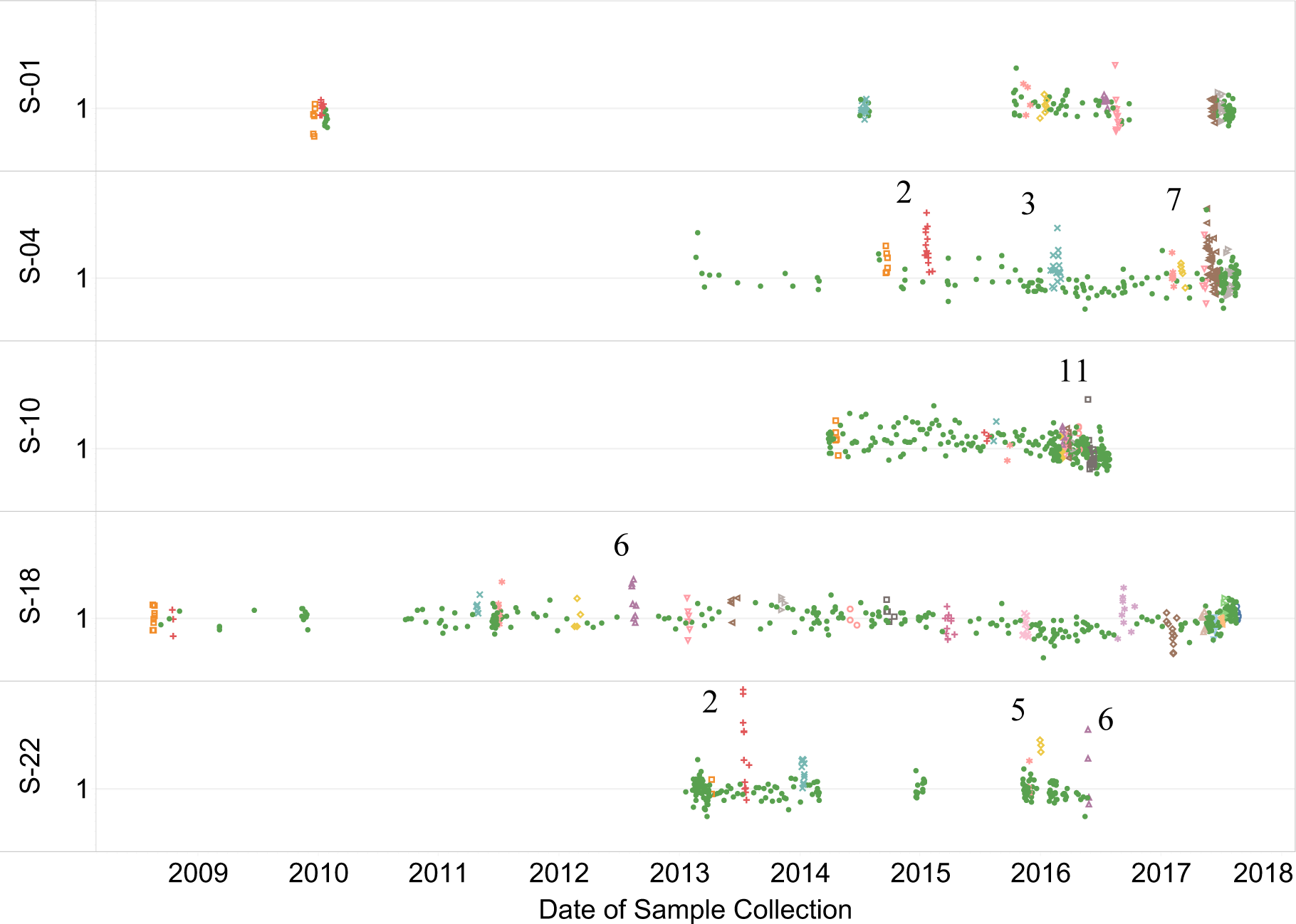
Values for MPO normalized by personal baseline showing several infection events in which MPO temporarily increased.

**Supplementary Fig 4.**
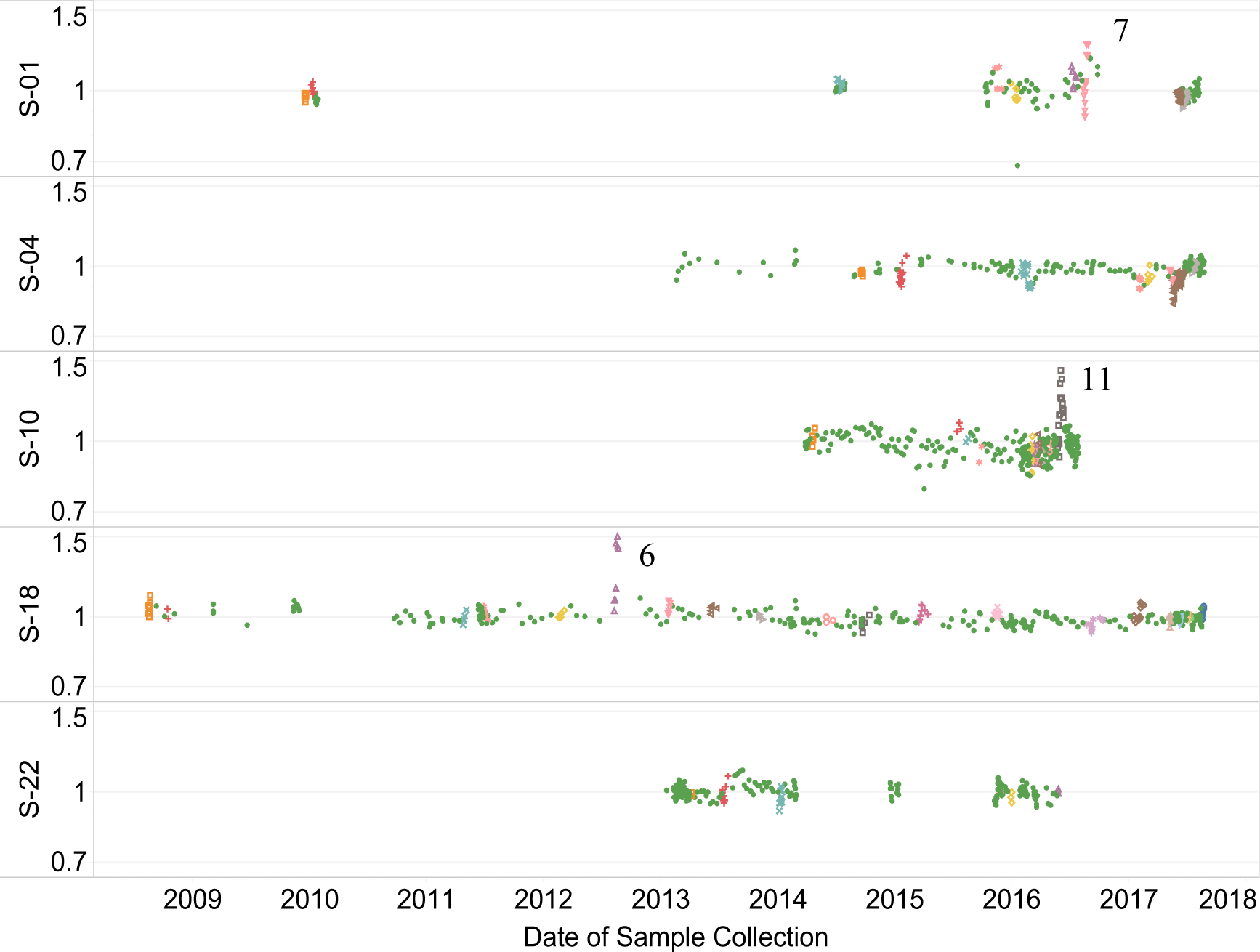
Values for IgM normalized by personal baseline showing two infection events (S-10 E11 and S-18 E6) in which IgM temporarily increased by more than 50% and the returned to baseline levels. A third event (S-01 E7) showed a temporary increase of about 30%.

## Abbreviations

A1AG: α-1-acid glycoprotein
Alb: Albumin
C3: Complement C3
CRP: C-reactive protein
FibG: Fibrinogen γ chain
Hp: Haptoglobin
Hx: Hemopexin
IgM: Immunoglobulin M
LPSBP: LPS-binding protein
MBL: Mannose binding lectin
MPO: Myeloperoxidase
SAA: Serum amyloid A
APR: acute phase response
SISCAPA: Stable Isotope Standards and Capture by Anti-Peptide Antibodies
PAR: peak area ratio
MRM-MS: multiple reaction monitoring MS

